# Seroconversion following SARS-CoV-2 Infection or Vaccination in Pediatric IBD Patients

**DOI:** 10.1101/2021.05.18.21257400

**Authors:** Elizabeth A. Spencer, Eyal Klang, Michael Dolinger, Nanci Pittman, Marla C. Dubinsky

## Abstract

**Objective:** Inflammatory bowel disease (IBD) patients are commonly treated with immunomodulatory medications, and the effect of these medications on seroconversion to SARS-CoV-2 infection and vaccination are scant, particularly in pediatrics. We sought to determine serologic responses to SARS-CoV-2 infection and vaccination in pediatric IBD patients.

**Design:** We conducted a single-center, retrospective study of all pediatric (≤21 years old) IBD patients in whom a SARS-CoV-2 IgG Antibody Assay was performed between April 2020 and May 2021 at our tertiary care center. This assay measures IgG antibody to the full-length SARS-CoV-2 spike protein and was routinely collected at infusion and outpatient clinic visits. The primary outcome was SARS-CoV-2 seroconversion, and the secondary outcome was titer level, with high titer defined as ≥960 titer or >40 AU/mL. Clinical characteristics, including demographics, IBD location, behavior, activity, and therapy, SARS-CoV-2 exposures, COVID-19 testing and symptoms, SARS-CoV-2 infection status (WHO COVID-19: Case Definitions, 2020) and COVID-19 vaccination status and type, were gathered, and univariate analyses examined associations between clinical characteristics and outcome measures.

**Results:** There were 340 pediatric patients with SARS-CoV-2 Antibody Testing; 15% for confirmed or probable COVID-19, 2% for suspected COVID-19, 16% for asymptomatic exposure to a close contact with SARS-CoV-2 infection, 61% without any prior symptoms or exposures, and 6% for history of COVID-19 vaccination. Patients with confirmed or probable COVID-19 infection had a 90% rate of seroconversion, with 76% of these patients on biologic therapy. Patients post-infection without seroconversion had a significantly longer interval between infection and antibody assay (*P*=0.03). Within those with asymptomatic SARS-CoV-2 exposure, 43% had seroconversion, and there were no identified clinical characteristics associated with positive titer.

All pediatric patients who received vaccination seroconverted, and all who received mRNA vaccinations, including one after a single dose, achieved high titer levels; 100% of those who received vaccination were on biologic or small molecule therapy, including one on combination therapy with ustekinumab and tofacitinib.

**Conclusion:** Pediatric IBD patients have strong serologic antibody responses to SARS-CoV-2 infection and COVID-19 vaccination despite high rates of immunomodulatory therapy.

## Introduction

Protective immunity to SARS-CoV-2, either naturally-induced by infection or artificially-induced or augmented by vaccination, is vital to reducing the transmission of SARS-CoV-2 and the burden of severe COVID-19. Data on the impact of immunomodulatory therapies used to treat inflammatory bowel disease (IBD) on both types of protective immunity have been scant, with initial reports of attenuation of natural immunity and vaccine efficacy by TNF antagonists^1,2^. However, two doses of COVID-19 vaccine or a single dose in those with a history of SARS-CoV-2 infection have both been shown to induce seroconversion in a large majority of patients^2,3^. There is little data on seroconversion in pediatric IBD, a population that may have distinct immunologic responses to SARS-CoV-2, given that increased age has been associated with lower antibody concentrations post-COVID-19-vaccination^2^. Understanding seroconversion is particularly important as vaccination becomes more widely available in pediatrics with the recent emergency use approval of BNT162b2 (Pfizer-BioNTech) down to 12 years of age^4^. We, therefore, sought to evaluate and compare serologic responses to SARS-CoV-2 infection and vaccination in a pediatric IBD cohort.

## Methods

We conducted a retrospective chart review of all IBD patients ≤21 years old in whom a SARS-CoV-2 IgG Antibody Assay was performed between April 2020 and May 2021 at our tertiary care center. The COVID-SeroKlir (Kantaro Biosciences, LLC, New York, NY) semi-quantitative SARS-CoV-2 IgG Antibody Assay, an ELISA measuring IgG antibody to the full-length SARS-CoV-2 spike protein with emergency use authorization, was routinely collected at infusion and outpatient clinic visits^5^. The study was approved by the Mount Sinai Institutional Review Board.

Electronic medical records were reviewed, and data were collected on demographics, IBD location and behavior (Paris classification)^6^, exposure(s) to SARS-CoV-2, and history of SARS-CoV-2 infection/symptoms. Indication for antibody testing and titer levels, which were described as high titer or strongly positive (≥960 titer or >40 AU/mL), moderately positive (320-960 titer or 16-39 AU/mL), weakly positive (80-160 titer or 5-15 AU/mL), and negative,^5^ were recorded. Patients meeting WHO criteria for confirmed (laboratory confirmation of COVID-19 infection) or probable COVID-19 (either meeting defined clinical criteria with contact with a probable or confirmed case of COVID-19 *or* recent onset of anosmia/ageusia without an identified cause) were classified as having prior COVID-19 infection^7^.

Standard descriptive statistics, including frequency for categorical variables and median [interquartile range (IQR)] for continuous variables, were calculated unless otherwise stated. Univariate analyses were performed using Fisher’s Exact and Chi-Square for categorical variables and Mann-Whitney and Spearman’s Rank Coefficient for continuous variables where appropriate. Statistical analyses were performed using R 3.6.3 (The R Foundation for Statistical Computing, 2018) and SAS OnDemand for Academics 3.8 (SAS Institute Inc., Cary, NC, 2020). P-value ≤0.05 was considered statistically significant.

## Results

One or more SARS-CoV-2 IgG Antibody Assay(s) was performed in 340 pediatric patients; 15% for confirmed or probable COVID-19, 2% for suspected COVID-19, 16% for an exposure to a close contact with SARS-CoV-2 infection without clinical symptoms, 61% without any prior symptoms or exposures, and 6% for history of COVID-19 vaccination. In the 51 patients with confirmed or probable COVID-19, 90% had seroconversion (**Supplementary Table 1**). Of the five patients without evidence of seroconversion, three had confirmed, and two had probable COVID-19. Those that did not seroconvert were similar in age to the rest of the post-infection cohort (Median 20 [17-20] years, *P*-value= 0.15) with a trend to a higher proportion of patients with UC/IBD-U (60%, *P*-value=0.07); four (80%) patients were on a biologic therapy (3 infliximab, 1 ustekinumab) and the remaining patient was on 5-aminosalicylate (5-ASA). There was a significantly longer time interval between infection and titer level measurement in those with a negative titer (Negative: 257 [167-340] days; Positive: 112 [41-180] days; *P*-value: 0.03); however, titer level was not correlated with time from infection in the entire post-infection cohort (ρ = -0.06, *P*-value = 0.74).

Within the 16% with exposure to SARS-CoV-2 without clinical symptoms, 23 (43%) had a positive SARS-CoV-2 Antibody Assay. There were no identified clinical characteristics associated with seroconversion in the group exposed to SAR-CoV-2 without clinical symptoms (**Supplementary Table 2**).

Twenty patients had antibody testing after vaccination (**Table 1**). All patients seroconverted following vaccination, and all patients receiving mRNA vaccination had high titer levels. The patient who received a single dose of JNJ-78436735 (Johnson & Johnson) had a moderate titer level. All but one (95%) of those receiving a two-series mRNA vaccination had completed both vaccinations in the series; the single patient with an assay performed after only one dose of BNT162b2, with no prior history of SARS-CoV-2 infection, seroconverted. Titer level was not significantly associated with type of biologic or small molecule therapy (**Figure 1**); patients receiving mRNA-1273 (NIH-Moderna) did have significantly higher titer levels compared to BNT162b2 and JNJ-78436735 (P=0.005).

**Figure 1.**
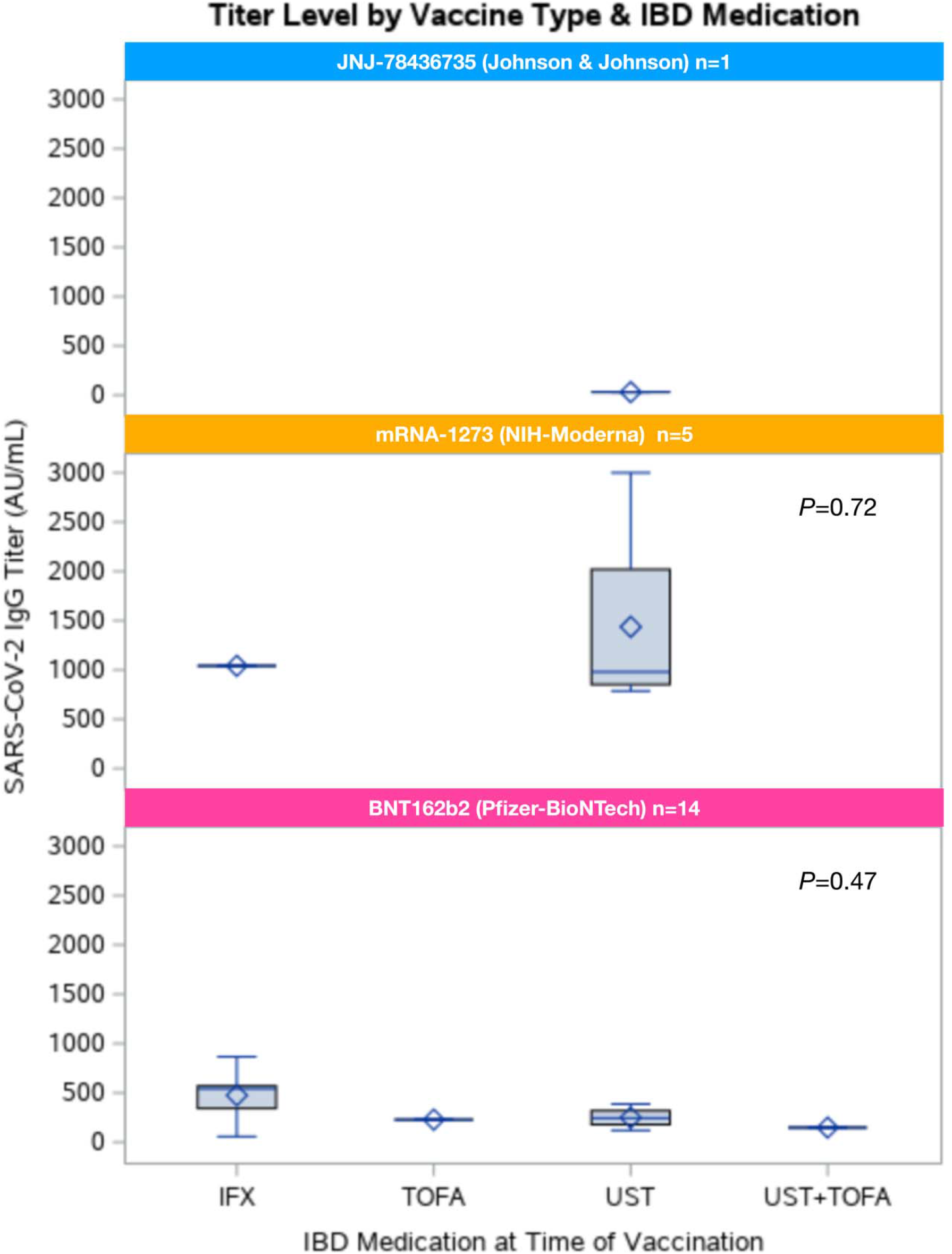
Titer Level by IBD Therapy and Vaccine Type Abbreviations: IFX, infliximab; TOFA, tofacitinib; UST, ustekinumab

**Table 1.**

| **Table 1** |  |
| --- | --- |
| **Clinical Characteristic** | **Vaccination +/- Prior Infection**<br>**n = 20** |
| **Demographics** |  |
| Male, N (%) | 12 (60) |
| Age (years), Median [IQR] | 18 [17-20] |
| **Vaccination Type** |  |
| BNT162b2 (Pfizer-BioNTech) | 14 (70) |
| mRNA-1273 (NIH-Moderna) | 5 (25) |
| JNJ-78436735 (Johnson & Johnson) | 1 (5) |
| **IBD Subtype, N (%)** |  |
| Crohn's Disease | 15 (75) |
| Ulcerative colitis | 5 (25) |
| **Age of Diagnosis, N (%)** |  |
| Diagnosis <17 years | 17 (85) |
| **Crohn's Disease Location** |  |
| Ileal | 4 (29) |
| Colonic | 1 (7) |
| Ileocolonic | 8 (57) |
| Isolated Upper Tract | 1 (7) |
| **Crohn's Disease Behavior, N (%)** |  |
| Non-penetrating, non-stricturing | 11 (85) |
| Stricturing | 1 (8) |
| Penetrating | 0 (0) |
| Stricturing & Penetrating | 1 (8) |
| Perianal Disease | 2 (15) |
| **Ulcerative Colitis/IBD-U, N (%)** |  |
| Proctitis | 0 (0) |
| Left-sided | 1 (20) |
| Extensive/Pancolitis | 4 (80) |
| **IBD Therapy, N (%) (Supplementary Figure 1B)** |  |
| Biologic Therapy | 19 (95) |
| Infliximab | 7 (37) |
| Adalimumab | 2 (11) |
| Ustekinumab | 10 (53) |
| Vedolizumab | 0 (0) |
| Tofacitinib | 2 (10)<sup>+</sup> |
| **Disease Activity, N (%)** |  |
| Clinical Remission* at Time of Vaccination | 17 (89) |
| **SARS-CoV-2 Antibody Testing** |  |
| Antibody positive, N (%) | 20 (100) |
| Median [IQR] Time from Last Vaccination to Titer (days) | 29 [14-37]<sup>^</sup> |
| High titer, N (%) | 18 (95)<sup>^</sup> |
| History of Infection, N (%) | 5 (25) |

\*Clinical Remission: Partial Mayo Score <2 or Harvey Bradshaw Index <4; <sup>^</sup>Single patient with qualitative titer only available; <sup>+</sup>One patient was on combination ustekinumab and tofacitinib

## Discussion

Herein we report robust serologic antibody responses to SARS-CoV-2 infection and COVID-19 vaccination in a pediatric IBD cohort. All of our patients seroconverted after vaccination even in the setting of biologic and small molecule usage, which was similar to the findings in an adult IBD study published out of our center^3^. Moreover, nearly all (90%) of our patients had seroconversion following SARS-CoV-2 infection, which was higher than the rates seen in Kennedy et al., suggesting improved post-infection seroconversion in pediatrics^1^. The high titer levels achieved in a large number of those who seroconverted are thought to confer protection; however, the association with elapsed time from SARS-CoV-2 exposure to negative level warrants continued investigation into the longevity of the protection conferred as well as a more detailed cataloging of the complexities of the immunoprotective response beyond IgG antibodies. While we are limited by our small sample size and variable times to assay, this study provides important reassurances to pediatric gastroenterologists, patients, and families and lends further support to expert consensus recommendations for vaccination of IBD patients^8^.

## Data Availability

Data archived in RedCap Database at our institution.

## Acknowledgements

The authors wish to thank the pediatric gastroenterologists at the Mount Sinai IBD Center and Randa Samaha, FNP.

---

**Supplementary Table 1.**

| **Clinical Characteristic** | **SARS-CoV-2 Infection without**<br>**Vaccination**<br>**n = 51** |
| --- | --- |
| **Demographics** |  |
| Male, N (%) | 29 (57) |
| Age (years), Median [IQR] | 16 [13-18] |
| **IBD Subtype, N (%)** |  |
| Crohn's Disease | 39 (76) |
| Ulcerative colitis | 9 (18) |
| IBD-Unspecified (IBD-U) | 3 (6) |
| **Age of Diagnosis, N (%)** |  |
| Diagnosis <17 years | 46 (90) |
| **Crohn's Disease Location** |  |
| Ileal | 10 (26) |
| Colonic | 2 (5) |
| Ileocolonic | 26 (67) |
| Isolated Upper Tract | 1 (3) |
| **Crohn's Disease Behavior, N (%)** |  |
| Non-penetrating, non-stricturing | 36 (92) |
| Stricturing | 1 (3) |
| Penetrating | 1 (3) |
| Stricturing & Penetrating | 1 (3) |
| Perianal Disease | 5 (13) |
| **Ulcerative Colitis/IBD-U, N (%)** |  |
| Proctitis | 2 (17) |
| Left-sided | 1 (8) |
| Extensive/Pancolitis | 9 (75) |
| **IBD Therapy, N (%) (Supplementary Figure 1A)** |  |
| Biologic Therapy | 39 (76) |
| Infliximab | 17 (44) |
| Adalimumab | 9 (23) |
| Ustekinumab | 10 (26) |
| Vedolizumab | 3 (8) |
| 5-ASA | 6 (12) |
| Antibiotics | 1 (2) |
| Tofacitinib | 0 (0) |
| No IBD medications | 7 (14) |
| **Disease Activity, N (%)** |  |
| Clinical Remission* at Time of Infection | 44 (86) |
| **SARS-CoV-2 Antibody Testing** |  |
| Antibody positive, N (%) | 46 (90) |
| Median [IQR] Time from Infection to Titer (days) | 117 [52-191] |
| High titer, N (%) | 20 (42) |

\*Clinical Remission: Partial Mayo Score <2 or Harvey Bradshaw Index <4

**Supplementary Table 2.**
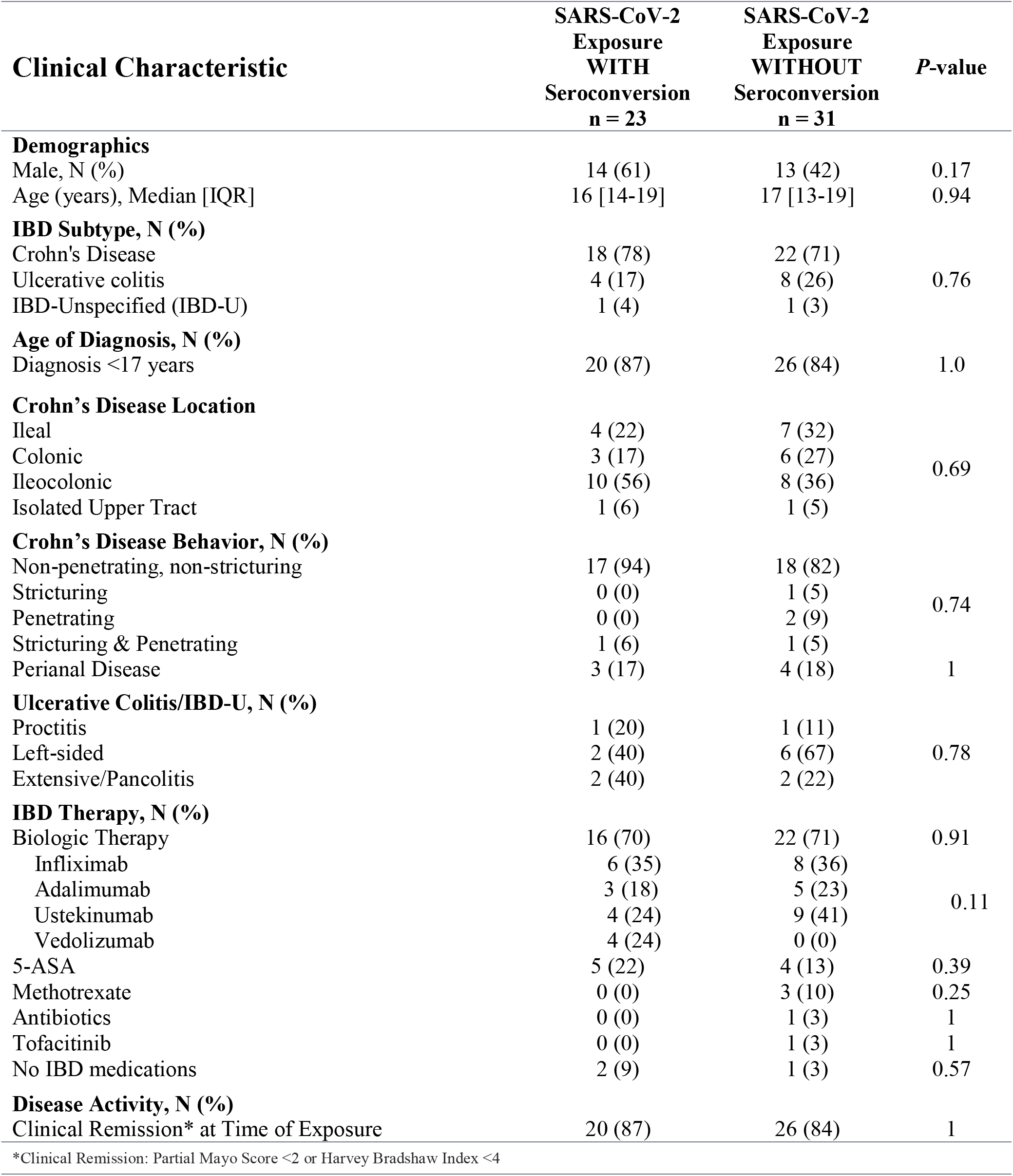

**Supplementary Figure 1A.**
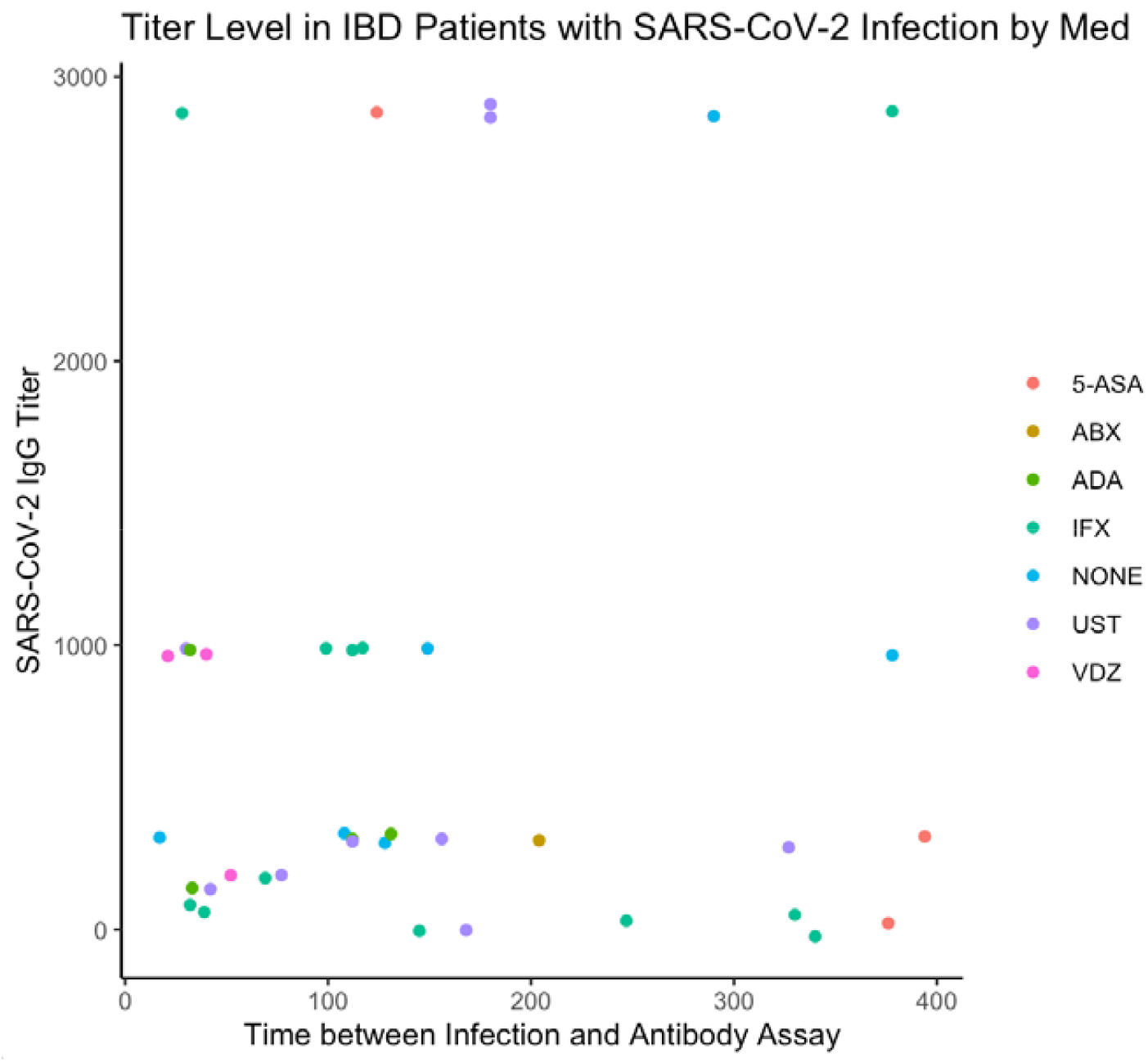
Titer Level in Pediatric IBD Patients with SARS-CoV-2 Infection by Medication Abbreviations: 5-ASA, 5-aminosalicylate; ABX, antibiotics (Ciprofloxacin/Metronidazole); ADA, adalimumab; IFX, infliximab; NONE, no IBD medications; UST, ustekinumab; VDZ, vedolizumab

**Supplementary Figure 1B.**
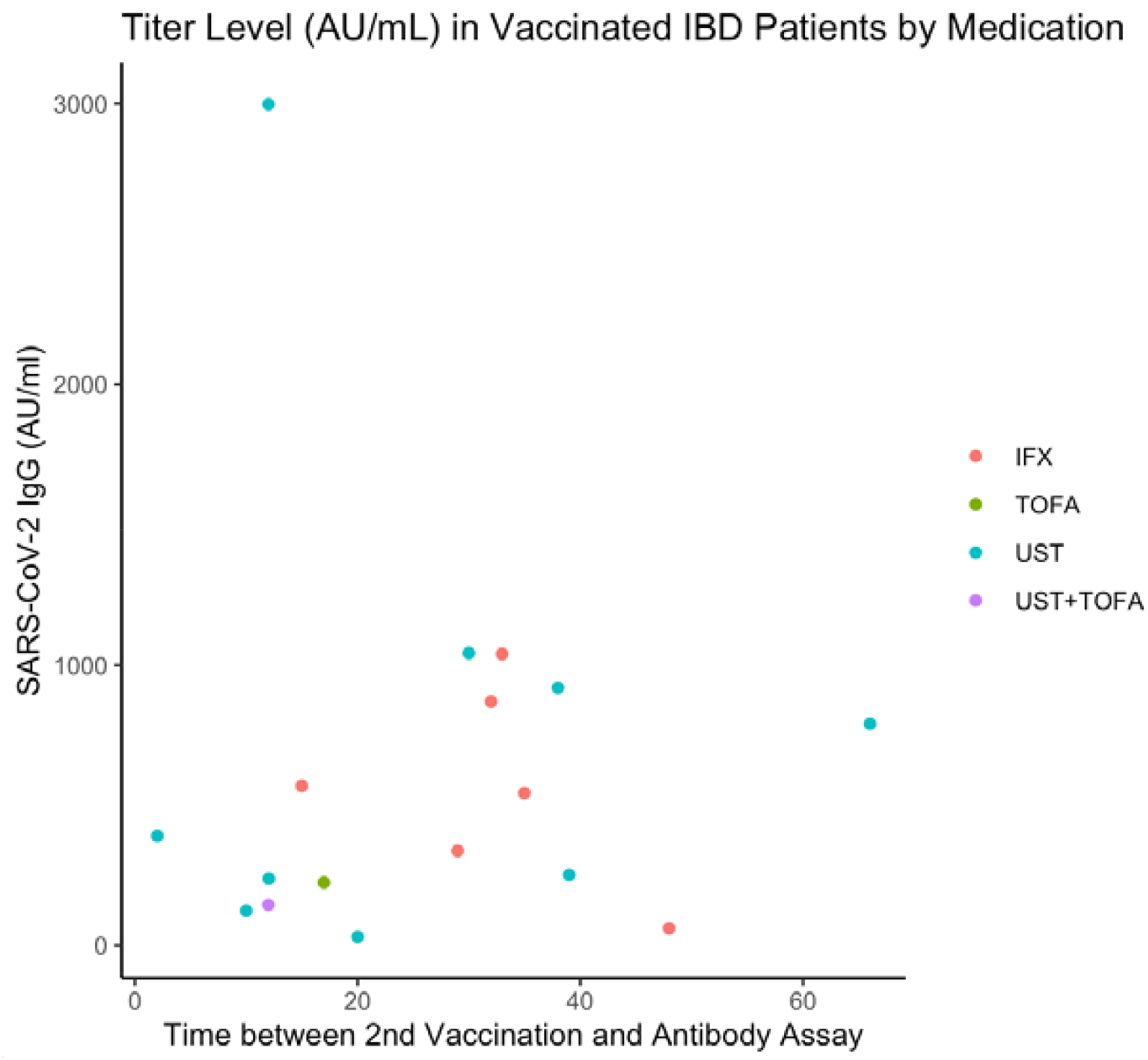
Titer Level (AU/mL) in Vaccinated Pediatric IBD Patients by Medication Abbreviations: IFX, infliximab; TOFA, tofacitinib; UST, ustekinumab

